# Associations of SARS-CoV-2 serum IgG with occupation and demographics of military personnel

**DOI:** 10.1101/2021.04.21.21255881

**Authors:** Joseph Zell, Jon Klein, Carolina Lucas, Martin Slade, Jian Liu, Akiko Iwasaki, Adam V Wisnewski, Carrie A Redlich

## Abstract

**Background:** Countries across the globe have mobilized their armed forces in response to COVID-19, placing them at increased risk for viral exposure. Humoral responses to SARS-CoV-2 among military personnel serve as biomarkers of infection and provide a basis for disease surveillance and recognition of work-related risk factors.

**Methods:** Enzyme-linked immunosorbent assays (ELISA) were used to measure SARS-CoV-2 spike antigen-specific IgG in serum obtained from N=995 US National Guard soldiers between April-June 2020. Occupational information, e.g. military operating specialty (MOS) codes, and demographic data were obtained via questionnaire. Plaque assays with live SARS-CoV-2 were used to assess serum neutralizing capacity for limited subjects (N=12).

**Results:** The SARS-CoV-2 IgG seropositivity rate among the study population was 10.3% and significantly associated with occupation and demographics. Odds ratios were highest for those working in MOS 2T-Transportation (3.6; 95% CI 0.7-18) and 92F-Fuel specialist/ground and aircraft (6.8; 95% CI 1.5-30), as well as black race (2.2; 95% CI 1.2-4.1), household size ≥6 (2.5; 95% CI 1.3-4.6) and known COVID-19 exposure (2.0; 95% CI 1.2-3.3). Seropositivity tracked along major interstate highways and clustered near the international airport and the New York City border. SARS-CoV-2 spike IgG^+^ serum exhibited low to moderate SARS-CoV-2 neutralizing capacity with IC_50s_ ranging from 1:15 to 1:280. In limited follow-up testing SARS-CoV-2 serum IgG levels remained elevated up to 7 months.

**Conclusions:** The data highlight increased SARS-CoV-2 seroprevalence among National Guard vs. the local civilian population in association with transportation-related occupations and specific demographics.

## INTRODUCTION

The armed forces of most countries play an important role in national/local disasters and emergencies as well as in response to military threats. Countries across the globe have mobilized their armed forces in response to the COVID-19 pandemic, placing them at increased risk for viral exposure (1-3). COVID-19 outbreaks among military personnel highlight their susceptibility to SARS-CoV-2 infection and the importance of disease surveillance of this critical workforce (4-8).

The National Guard of United States (US) Military is the division responsible for protecting civilians in the homeland, with each state/territory maintaining its own troops. To assist in the COVID-19 pandemic, governors across all 50 states, Puerto Rico, Guam, the U.S. Virgin Islands, and Washington D.C. deployed their local Army/Air National Guard Units (9). At the height of deployment levels in May 2020 nearly 50,000 troops were activated for missions that included sorting and delivering medical supplies, cleaning high-traffic public spaces, food distribution, and assisting nursing homes with testing, cleaning, and caring for patients. The deadline for National Guard deployments originally set to end Aug. 21, 2020 was extended through March 31, 2021 in forty-eight states and three territories (10).

In Connecticut, USA, a state where SARS-CoV-2 was detected early during the pandemic, the National Guard was at the forefront of anti-COVID-19 efforts. To help ensure workforce protection, the state’s military leadership adopted a proactive stance toward surveillance via serology. Data from N=955 soldiers, airmen, and militia members to undergo COVID-19 antibody testing during April-June 2020 are analyzed here.

The present study results provide insight into the prevalence and localized distribution of COVID-19 cases early in the pandemic. The data identify significant associations of viral-specific serum IgG with demographics as well as job title as defined by military occupational specialty (MOS) code. MOS codes (usually starting with a specific number-letter combination) are used by the US military to define jobs with similar duties and functions. We further evaluated viral neutralizing capacity of serum from a subset of subjects and the duration of SARS-CoV-2 serum IgG in limited follow-up samples available up to 7 months after the initial test. The significance of the present data and implications for COVID-19 and future pandemics are discussed.

## MATERIALS AND METHODS

### Human subjects

Volunteers from the CT Army and Air National Guard were recruited to have their SARS-CoV-2 spike antigen-specific antibody levels tested. Subjects provided 3cc of blood by venipuncture using vacutainer tubes (Becton Dickinson; Franklin Lake, NJ); serum was separated and stored frozen at −80°C until tested by enzyme linked immunosorbent assay (ELISA). The studies were reviewed and ethical approval was given by the Yale University Institutional Review Board. All participants provided informed verbal consent.

### ELISA methods

ELISAs were performed as previously described with minor modifications (11, 12). In short, Triton X-100 and RNase A were added to serum samples at final concentrations of 0.5% and 0.5mg/ml respectively and incubated at room temperature (RT) for 30 minutes before use to reduce risk from any potential virus in serum. 96-well MaxiSorp plates (ThermoFisher, Waltham, MA) were coated with 50 μL/well of recombinant SARS Cov-2 ectodomain (Sino Biologicals; Wayne, PA) at a concentration of 1 μg/ml in NaCO_3_ buffer pH 9.6 and incubated overnight at 4°C. The coating buffer was removed, and plates were incubated for 1h at RT with 200 μL of blocking solution (PBS with 3% milk powder). Serum was diluted 1:100 in dilution solution (PBS with 0.05% Tween20, 1% milk powder) and 100 μL of diluted serum was added for one hours at RT. Plates were washed three times with PBS-T (PBS with 0.1% Tween-20) and 50 μL of HRP anti-Human IgG Antibody (Pharmingen/BD Biosiences, San Jose, CA) or HRP anti-human IgA (BioLegend, San Diego, CA) were added at 1:2000-fold dilution. After 1 h of incubation at RT, plates were washed three times with PBS-T. Plates were developed with 100 μL of TMB Substrate Reagent Set (BD Biosciences, San Jose, CA) and the reaction was stopped when an internal pooled serum positive control sample reaches an OD of 1.0 at 650 nm, by the addition of 2 N sulfuric acid. Plates were then read at a wavelength of 450 nm with reference wavelength calibration (650nm).

### Cell lines and virus

VeroE6 kidney epithelial cells from ATTC were cultured in Dulbecco’s Modified Eagle Medium (DMEM) supplemented with 1% sodium pyruvate (NEAA) and 5% fetal bovine serum (FBS) at 37°C and 5% CO_2_. SARS-CoV-2, strain USA-WA1/2020, was obtained from BEI Resources (#NR-52281) and amplified in VeroE6 cells. Cells were infected at a multiplicity of infection level of 0.01 for four three days to generate a working stock. Culture supernatant was clarified by centrifugation (450*g* × 5min) and filtered through a 0.45-micron filter. The pelleted virus was then resuspended in PBS then aliquoted for storage at −80°C. Viral titers were measured by standard plaque assay using Vero E6 cells. Briefly, 300 µL of serial fold virus dilutions were used to infect Vero E6 cells in MEM supplemented NaHCO_3_, 4% FBS 0.6% Avicel RC-581. Plaques were resolved at 48h post infection by fixing in 10% formaldehyde for 1h followed by with 0.5% crystal violet in 20% ethanol staining. Plates were rinsed in water to plaques enumeration. All experiments were performed in a biosafety level 3 with the Yale Environmental Health and Safety office approval.

### Neutralization assay

Study subject sera were heat treated for 30m at 56°C. Sixfold serially diluted plasma, from 1:3 to 1:2430 were incubated with SARS-CoV-2 for 1 h at 37°C. The mixture was subsequently incubated with Vero E6 cells in a 6-well plate for 1h. Then, cells were overlayed with MEM supplemented NaHCO_3_, 4% FBS 0.6% Avicel mixture. Plaques were resolved at 40h post infection by fixing in 10% formaldehyde for 1h followed by staining in 0.5% crystal violet. All experiments were performed in parallel with negative controls sera, in a establish viral concentration to generate 120-150 plaques/well.

### Statistical Methods

Descriptive statistics (means and standard deviations for continuous variables and percentages for categorical variables) of both predictor and outcome variables for the entire study population were calculated. Linear regression models were developed to model the loge of spike antigen IgG response level as the data had a log normal distribution. Unadjusted (bivariate) models were developed for each predictor. Additionally, a parsimonious model was determined utilizing multivariate regression modeling incorporating a backward selection strategy with significance level-to-stay specified as α=0.05. To evaluate whether the spike antigen IgG response level exceeded a specified threshold, logistic regression models we utilized. Again, unadjusted (bivariate) models were developed for each predictor. Additionally, a parsimonious model was determined utilizing multivariate regression modeling incorporating a backward selection strategy with significance level-to-stay specified as α=0.05. SAS v9.4® was utilized for all statistical analyses.

## RESULTS

### Basic Demographics

This study included N=995 soldiers enlisted in the Connecticut Army and Air National Guard (Table 1). The racial composition was 7.7% African-Americans, 2.5% Asian, 3.9% mixed, and 72.5% white individuals. Race data was not available for N=120 (12.1%) individuals who provided only information on ethnicity (Hispanic). The study group’s age distribution was slightly shifted to the left compared to the working civilian population in CT (13), and contained predominately younger individuals (Supplemental Materials Figure S1). Similar to the overall US military population the study group was largely (79.2%) male and healthy (14). None of the participants were hospitalized with COVID-19 like illness since the onset of the SARS-CoV-2 pandemic.

**Table 1.**
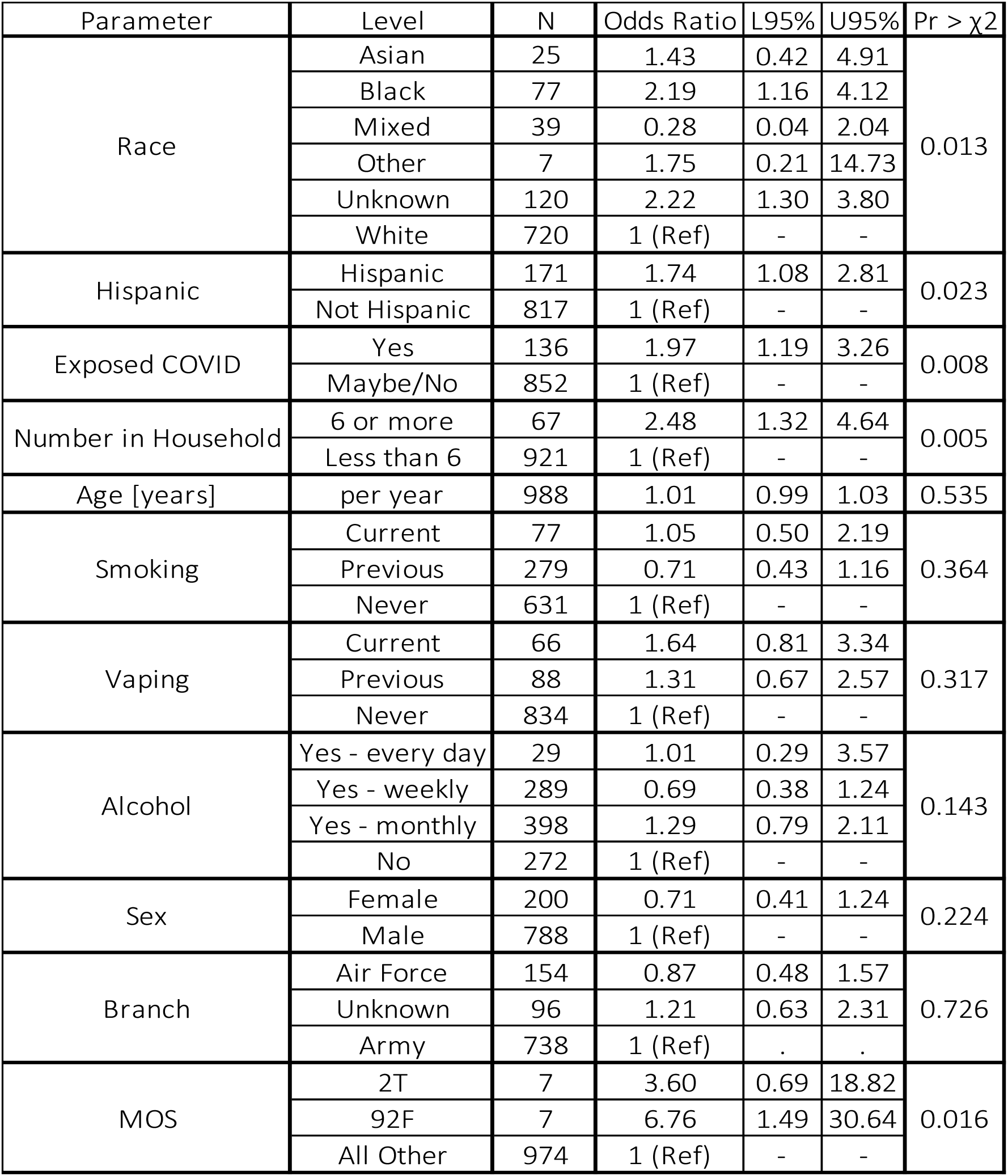
Demographic/Occupational Data: Associations with SARS-CoV-2 IgG.

| Parameter | Level | N | Odds Ratio | L95% | U95% | Pr > $\chi^2$ |
| --- | --- | --- | --- | --- | --- | --- |
| Race | Asian | 25 | 1.43 | 0.42 | 4.91 | 0.013 |
|  | Black | 77 | 2.19 | 1.16 | 4.12 |  |
|  | Mixed | 39 | 0.28 | 0.04 | 2.04 |  |
|  | Other | 7 | 1.75 | 0.21 | 14.73 |  |
|  | Unknown | 120 | 2.22 | 1.30 | 3.80 |  |
|  | White | 720 | 1 (Ref) | - | - |  |
| Hispanic | Hispanic | 171 | 1.74 | 1.08 | 2.81 | 0.023 |
|  | Not Hispanic | 817 | 1 (Ref) | - | - |  |
| Exposed COVID | Yes | 136 | 1.97 | 1.19 | 3.26 | 0.008 |
|  | Maybe/No | 852 | 1 (Ref) | - | - |  |
| Number in Household | 6 or more | 67 | 2.48 | 1.32 | 4.64 | 0.005 |
|  | Less than 6 | 921 | 1 (Ref) | - | - |  |
| Age [years] | per year | 988 | 1.01 | 0.99 | 1.03 | 0.535 |
| Smoking | Current | 77 | 1.05 | 0.50 | 2.19 | 0.364 |
|  | Previous | 279 | 0.71 | 0.43 | 1.16 |  |
|  | Never | 631 | 1 (Ref) | - | - |  |
| Vaping | Current | 66 | 1.64 | 0.81 | 3.34 | 0.317 |
|  | Previous | 88 | 1.31 | 0.67 | 2.57 |  |
|  | Never | 834 | 1 (Ref) | - | - |  |
| Alcohol | Yes - every day | 29 | 1.01 | 0.29 | 3.57 | 0.143 |
|  | Yes - weekly | 289 | 0.69 | 0.38 | 1.24 |  |
|  | Yes - monthly | 398 | 1.29 | 0.79 | 2.11 |  |
|  | No | 272 | 1 (Ref) | - | - |  |
| Sex | Female | 200 | 0.71 | 0.41 | 1.24 | 0.224 |
|  | Male | 788 | 1 (Ref) | - | - |  |
| Branch | Air Force | 154 | 0.87 | 0.48 | 1.57 | 0.726 |
|  | Unknown | 96 | 1.21 | 0.63 | 2.31 |  |
|  | Army | 738 | 1 (Ref) | . | . |  |
| MOS | 2T | 7 | 3.60 | 0.69 | 18.82 | 0.016 |
|  | 92F | 7 | 6.76 | 1.49 | 30.64 |  |
|  | All Other | 974 | 1 (Ref) | - | - |  |

### SARS-CoV-2 Serology Results and Demographic Associations

A total of N=103 (10.3%) of the subjects tested positive for SARS-CoV-2 spike antigen serum IgG, a rate 2.5-fold higher than the regional population at that time (15). Unadjusted logistic regression models (Table 1) demonstrated significant (*p* < 0.024) associations of SARS-CoV-2 IgG positivity with race and ethnicity, with notably increased odd ratios for black and Hispanic individuals (OR 2.1, 95% CI 1.2-4.1, and OR 1.7; 95% CI 1.1-2.8). Significant association of SARS-CoV-2 IgG positivity was also observed with household size ≥6 (*p* < 0.005) and known exposure to COVID-19 (*p* < 0.009), but not with specific lifestyle habits, tobacco use, vaping or alcohol consumption/frequency. In a parsimonious adjusted logistic regression model (Supplemental materials Table S1) SARS-CoV-2 IgG positivity remained significantly associated with race, household size and known exposure to COVID-19 (*p* < 0.017, 0.012, and 0.008 respectively).

Linear regression analysis of the data (supplemental Table S2) with SARS-CoV-2 IgG ELISA OD values quantitated as a continuous variable yielded stronger associations vs. logistic analysis with race, ethnicity, household size, and known COVID-19 exposure. Linear regression analysis further revealed a significant association of SARS-CoV-2 IgG levels with gender (*p* < 0.0002). While males were not more likely to be SARS-CoV-2 IgG positive than females, when they were positive, they had significantly (*p*<0.001) higher SARS-CoV-2 IgG ELISA OD values, on average by 0.25 OD units (Figure 1). Race, household size, known COVID-19 exposure, and gender remained significantly associated with SARS-CoV-2 IgG levels after parsimonious adjustment of linear regression models (Supplemental Materials Table S3).

**Figure 1.**
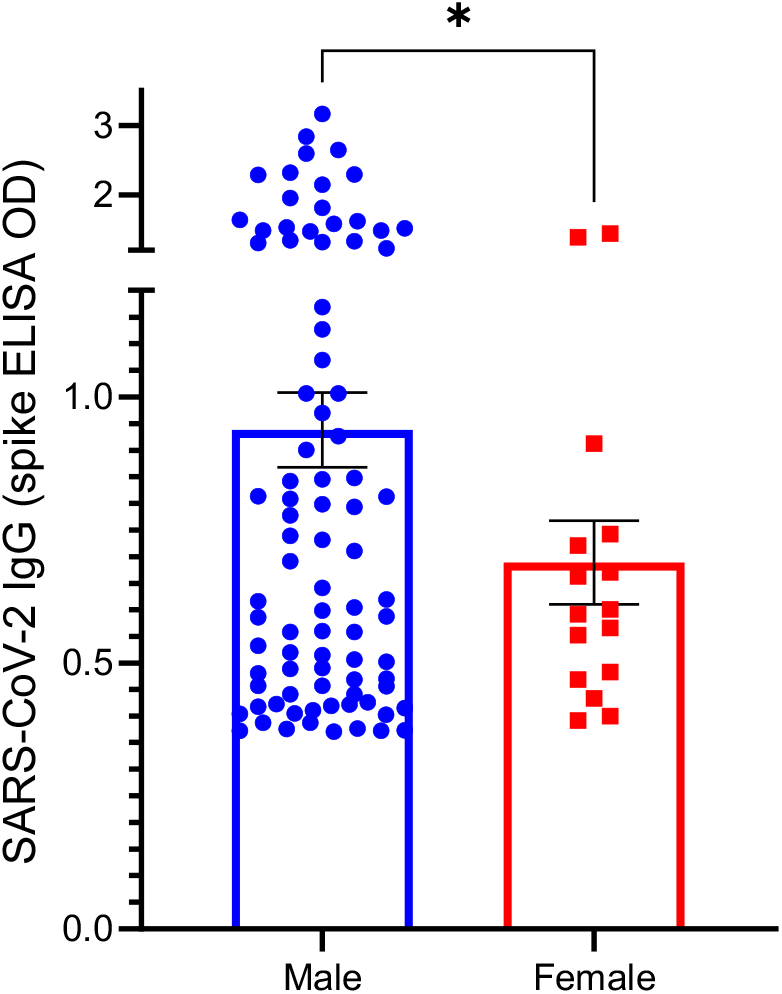
Spike antigen-specific serum IgG levels in males vs. females. The amount of spike antigen -specific IgG was quantitated based on ELISA OD values (Y-axis) and positive test results (OD > 0.356) are compared for males and females. * indicates significant (*p* < 0.023) difference between genders.

### SARS-CoV-2 Serology Results and Occupational Associations

We further evaluated potential associations of SARS-CoV-2 serum IgG with exposures that might be connected to work, using military operation specialty (MOS) codes to define job title and related duties. Greater than 2/3 of the troops belonged to MOS groups that contained N ≥ 7 people. Logistic and linear regression analysis demonstrated significant associations of SARS-CoV-2 IgG (p <0.016 and *p* < 0.002 respectively) with MOS codes 2T and 92F (Table 1 and Fig. 2). MOS 2T performs and manages activities related to transportation by ground and air, while MOS 92F is a fuel specialist for both ground and air transport. Troops with MOS codes 2T and 92T come in contact with individuals and cargo from around the US and the world via air transport. MOS 2T and 92F remained significantly associated with SARS-CoV-2 serum IgG levels after parsimonious adjustment of linear regression models, accounting for demographic associations (Supplemental Materials Table S3).

**Figure 2.**
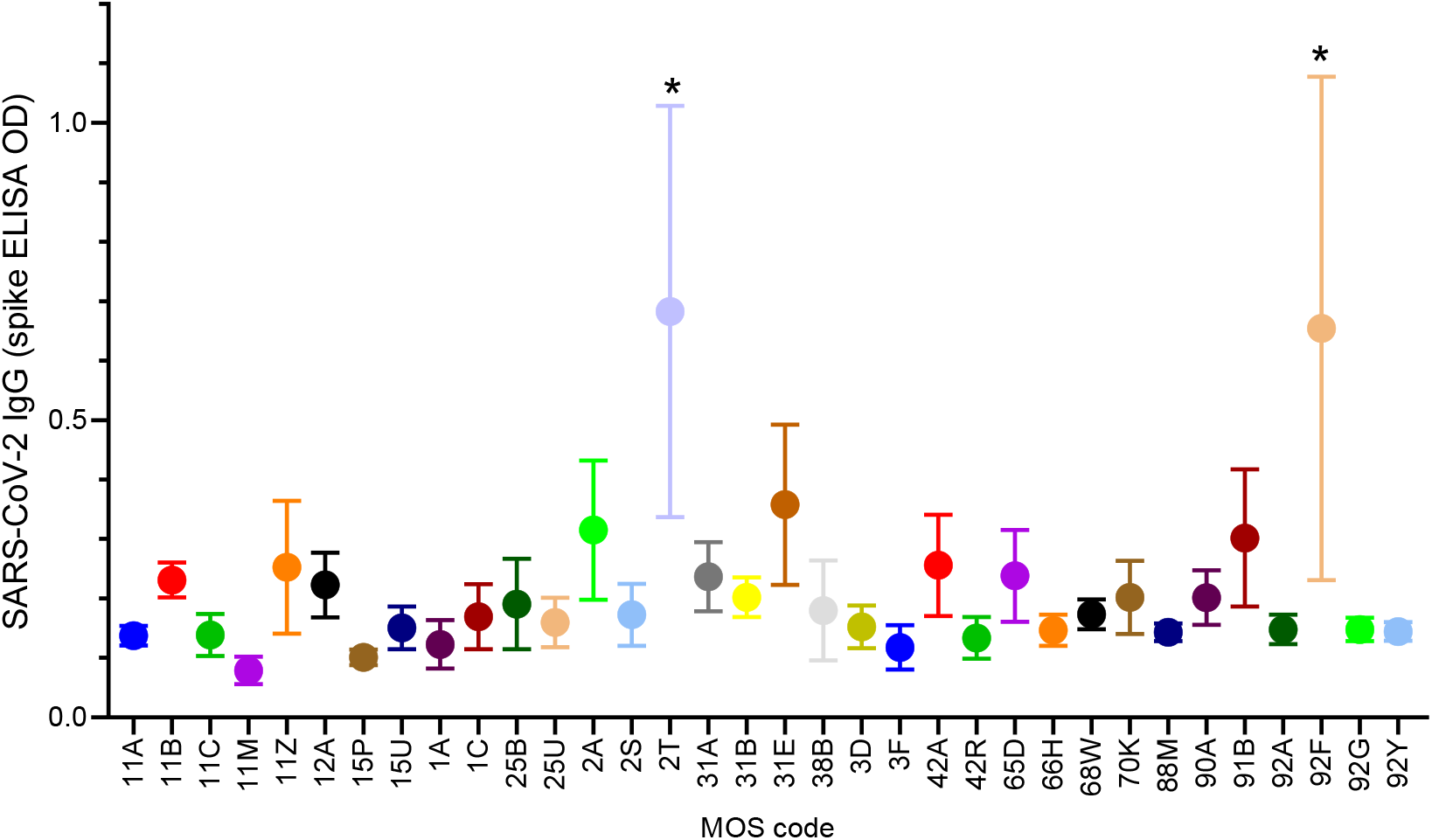
Spike antigen-specific IgG in serum from subjects with different MOS codes. The mean and standard error of spike IgG ELISA OD values are shown for subjects that belong to different MOS groups with N ≥ 7 per MOS. * indicates significant (*p* < 0.017) difference vs. other all other MOS codes.

Among those subjects that held MOS codes with N ≤6 persons it was notable that 2/2 subjects with MOS code 155E (C-12 fighter pilots) were SARS-CoV-2 IgG positive, as were several subjects with MOS codes for nursing subspecialties, MOS 66-68 (Supplemental Materials Fig S2). However, the low number of subjects with the corresponding MOS codes lacked statistical power to attribute significance to the observed associations with MOS 155E or 66-68.

### SARS-CoV-2 Serology Results and Geolocation

The distribution of SARS-CoV-2 IgG positivity throughout CT state was visualized based on zip code as shown in Figure 3. The notable hot spots were in Stamford, the 1^st^ major city north of New York City (NYC), the Northern part of the state near the international airport, and the zip codes for Bristol, Bridgeport and Waterbury; areas in CT’s upper quartile for population density and diversity, but lower quartile for median household and per capita income. The data are consistent with NYCs concomitant outbreak and the socio-economics of COVID-19, and further suggest association of infection with travel, given the number SARS-CoV-2 IgG positive cases near the airport/interstate highway, and the association with MOS codes 2T and 92F.

**Figure 3.**
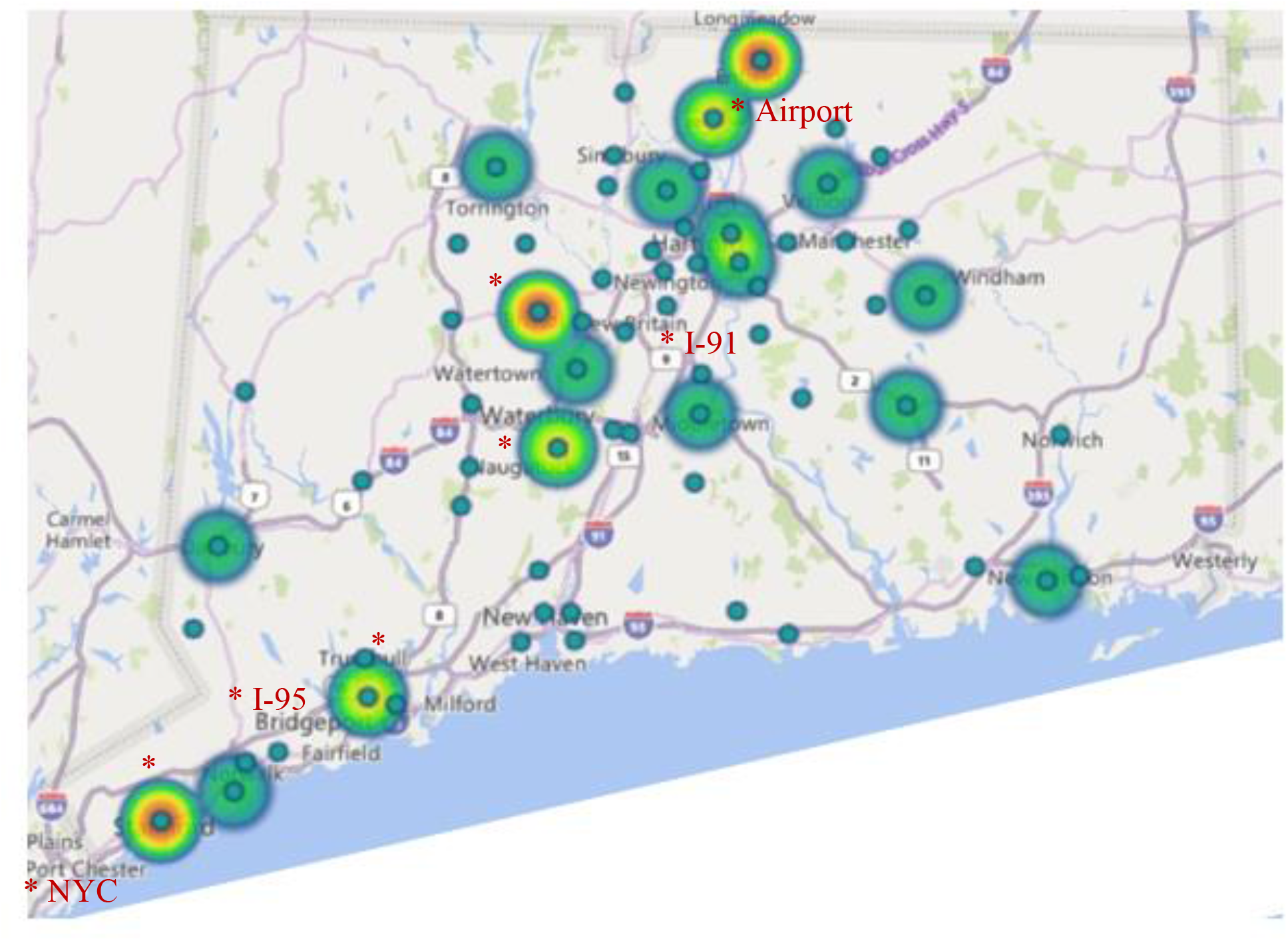
Geolocation of SARS-CoV-2 IgG+ subjects according to home zip code. The number of SARS-CoV-2 IgG+ subjects per zip code was mapped based on home zip code. Note highlighted hot spots (*) along the major (I-95 and I-91) interstate highways, Stamford, the town just north of NYC, Bristol, Bridgeport, Waterbury, and the north-central part of the state near the Hartford-Springfield International Airport.

### SARS-CoV-2 neutralization capacity of serum

SARS-CoV-2 serum neutralizing capacity of non-hospitalized individuals with mild or asymptomatic COVID-19 is relatively understudied vs. that of hospitalized patients with more severe disease (16). We measured the neutralizing ability of serum from a subgroup of subjects (N=12) with positive SARS-CoV-2 IgG test results in a live viral plaque assay. As shown in Fig. 4, all twelve subjects’ serum had significant neutralizing capacity with IC_50s_ ranging from 1:15 to 1:280. On average the subjects serum neutralizing capacity was comparable, albeit lower, than that of hospitalized patients as we recently published (17). Notably, serum spike IgG ELISA levels (OD) were closely related (*r*=.75) to the observed variability in IC_50_.

**Figure 4.**
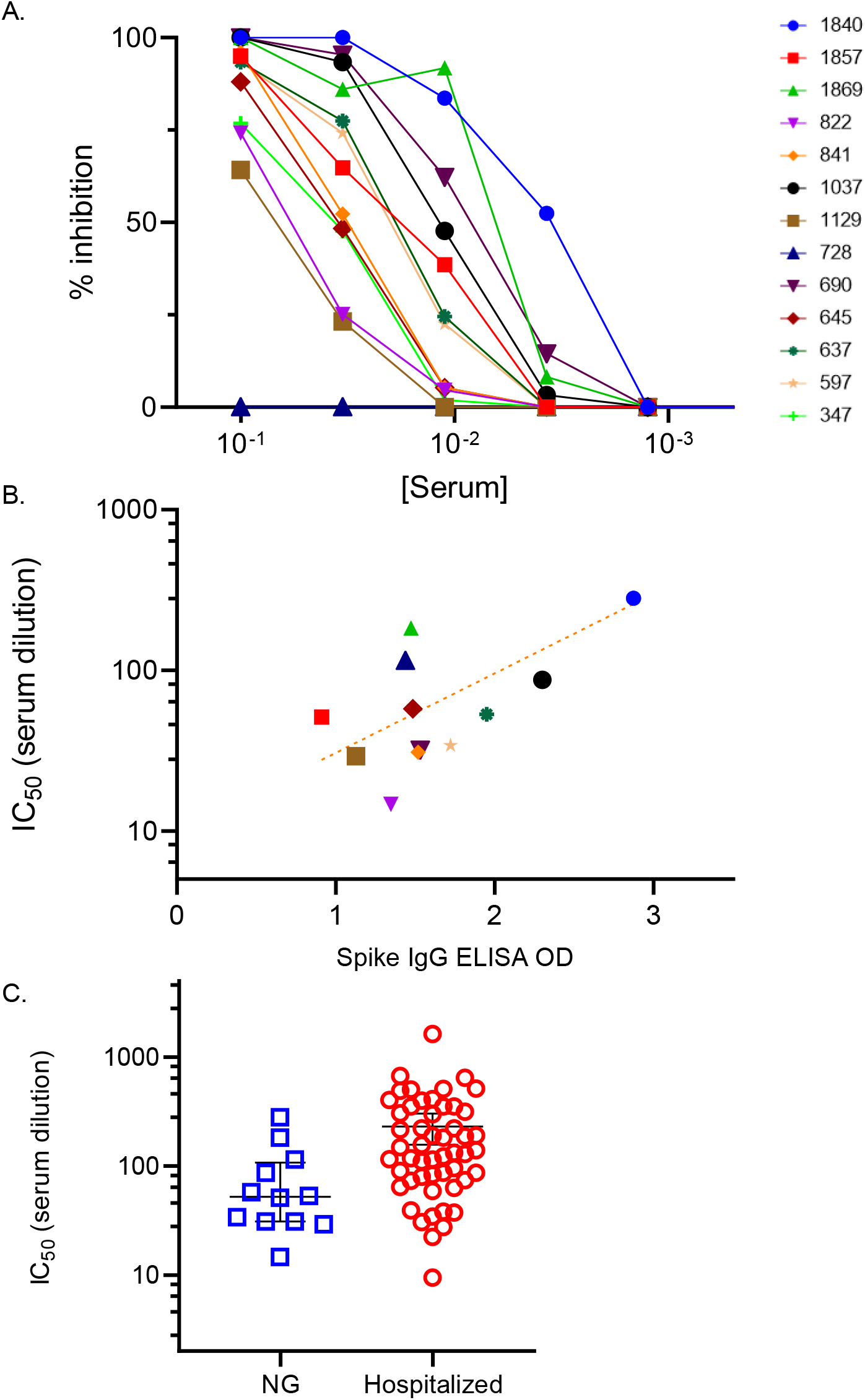
Neutralizing capacity of serum from individuals with mild/asymptomatic COVID-19, not requiring hospitalization. (A) Serum was titrated in a live viral plaque assay to measure neutralization capacity (% inhibition of plaque formation) expressed on the Y axis vs. serum concentration (X-axis). Different color line/symbol represents data from a single subject with ID# shown in key in upper right, and negative control (blue triangles). (B) Correlation between the IC_50_ (Y-axis) and the spike IgG ELISA OD values (X-axis). Each symbol represents the average of replicate data points from a single individual in key above. r^2^=.567 for the best fit trend line, log(y) = .495x + 0.991. (C) Comparison of serum neutralizing activity, IC_50_ (Y-axis) of the present National Guard troops (NG) vs. hospitalized study subjects recently published (17). Each symbol represents data from a single subject’s serum titration. The mean and standard error for the different groups of study subjects is shown.

### Longevity of SARS-CoV-2 serum IgG

Persistence of SARS-CoV-2 IgG in serum was evaluated in a limited number of subjects that returned for repeat testing. Follow-up samples obtained 6 weeks to 5 months after the initial test showed no significant difference in average SARS-CoV-2 spike IgG serum levels vs. the initial time point (Fig. 5). However, the lack of change may be obscured by differences in time of sample collection relative to infection, as some subjects SARS-CoV-2 spike IgG serum levels decreased, while others increased. Perhaps most notably, four subjects remained SARS-CoV-2 spike IgG positive for >7 months after the initial test, with sustained elevated ELISA signal.

**Figure 5.**
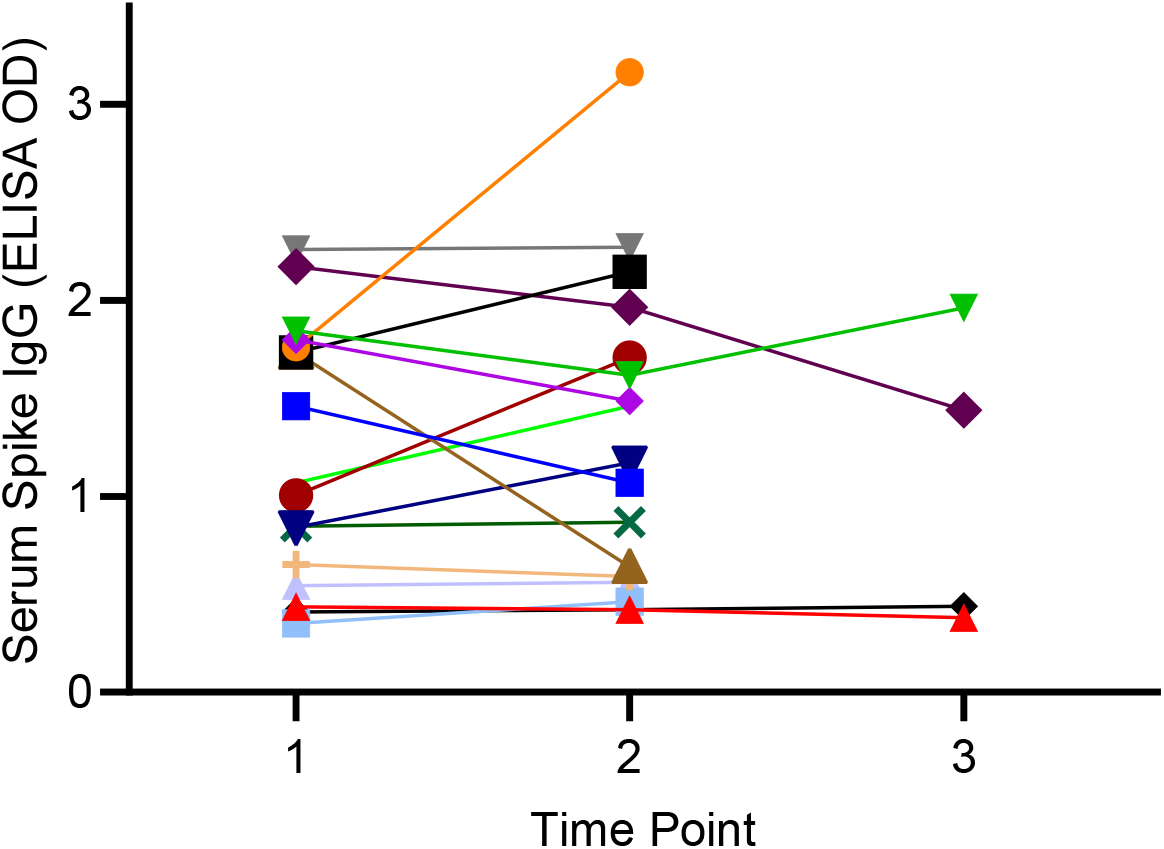
Longevity of serum SARS-CoV-2 spike IgG. The amount of spike antigen-specific IgG was measured based on the ELISA OD (Y-axis) over time (X-axis). Time point 1 = baseline, time point 2 = 6 weeks to 5 months, time point 3 = 7 months. Each different color symbol/line represents the mean of replicate data from a single time point for a single subject. The mean OD at time point 2 is not significantly different (*p* > 0.05) from time point 1.

## CONCLUSIONS

Sero-surveillance of US CT National Guard troops from April-June 2020 demonstrated a high prevalence (10.3%) of SARS-CoV-2 IgG compared to the local civilian population (4.1%) at the same time (15). SARS-CoV-2 spike antigen specific serum IgG was significantly associated with demographics and occupation, as defined by MOS code, and clustered in distinct locations within the state. Serum from a subset of individuals tested was found to possess viral neutralizing capacity with IC_50s_ between 1:15 and 1:280. In limited follow up samples, SARS-CoV-2 IgG serum levels remained elevated up to 7 months. Together the data demonstrate the development of protective adaptive humoral immune responses in working-age adults with mild/asymptomatic COVID-19 (not requiring hospitalization) and demonstrate the utility of sero-surveillance of healthy populations to identify at risk subgroups that might benefit from target intervention.

The occupational association of specific MOS codes (2T, 92F) with SARS-CoV-2 serum IgG, and the distribution of cases along major interstate highway and near the international airport, are consistent with ground and air travel as COVID-19 risk factors. MOS 2T is an Air Force MOS code for specialists in transport and vehicle management, while MOS 92F codes Army fuel supply specialists that interacts with individuals traveling across the country and the world. Further support for a link between air travel and COVID-19 is the observation that 2/2 C-12 (small commuter transport) pilots in the present study were SARS-CoV-2 IgG+. Together the data highlight transportation-related occupations as important SARS-CoV-2 risk factors, independent of demographics.

The association of spike serum IgG with transportation-related MOS codes fits with the geolocation data showing the highest number of SARS-CoV-2 IgG+ subjects near the international airport and along the major interstate highways. The geographic prevalence of SARS-CoV-2 seropositivity partially overlaps with, but was distinct from, locations of highest population density and/or percentage of National Guard troops. The geolocation data do not rule out potential for infections in less populated/unsampled areas but highlight geographical regions in the state where travel provides increased opportunity for viral transmission.

The present data are consistent with the high prevalence of SARS-CoV-2 among black, Hispanic, and socio-economically disadvantaged populations reported in numerous studies to date (18-21). The findings suggest that racial and ethnic COVID-19 risk factors for US military personnel are similar to the US civilian population. Recognition of these known racial and ethnic disparities is important for maintaining combat readiness and national security, in addition to equitable distribution of health care and preventative services.

An distinct aspect of the present study is the focus on immune responses in healthy non-hospitalized working age adults with mild or asymptomatic disease (vs. the pathologic response of hospitalized COVID-19 patients with more severe clinical course). In healthy individuals adaptive humoral responses to SARS-CoV-2 are likely part of effective immunity, yet remain less studied than those of hospitalized patients (16). The present data demonstrate the viral neutralizing capacity of serum from a limited number of non-hospitalized asymptomatic healthy individuals (albeit with lower IC_50s_ than hospitalized individuals) and demonstrate an association with spike antigen-specific IgG levels. Furthermore, in limited subjects for which follow-up serum samples were available, spike antigen specific IgG levels persisted and remain elevated up to 7 months. Thus, adaptive humoral immune responses in healthy working age adults with mild/asymptomatic COVID-19 serve as biomarkers of infection amenable to disease surveillance efforts and demonstrate an association with anti-viral activity.

The strengths and weaknesses of this study should be realized when interpreting the findings’ significance. Strengths include the large study population (N=995), available occupation (MOS code) and demographic information, and the ability to follow individuals over time. Limitations of the study included the small sample (N=12) size tested for viral neutralizing capacity and at the 7 month time point (N=4), due to resource and sample availability. There was overlap in the association of SARS-CoV-2 serum IgG with certain risk factors, however, occupational (MOS codes 2T, 92F) associations remained highly significant in parsimoniously adjusted logistic and linear regression models (Supplemental materials Tables S1 and S3).

In summary, serology of US National Guard troops deployed to help combat COVID-19 during April-June 2020 demonstrate increased prevalence of SARS-CoV-2 spike antigen-specific IgG compared to the local civilian population (15) in association with specific occupations and demographics. The results further demonstrate the neutralizing capacity of serum from subjects with mild/asymptomatic COVID-19, not requiring hospitalization, and persistence of spike antigen-specific IgG for up to 7 months in limited subjects. Together the data demonstrate the utility of SARS-CoV-2 sero-epidemiology to identify specific sub-populations of the US National Guard at risk for COVID-19. The finding may extend to civilian populations and military forces in other countries, and to other infectious or otherwise immunogenic agents.

## Data Availability

All data are available in the manuscript or in the supplemental materials.

## REFERENCES

1. Kalkman JP. Military crisis responses to covid-19. Journal of Contingencies and Crisis Management 2020:10.1111/1468-5973.12328.

2. Gad M, Kazibwe J, Quirk E, Gheorghe A, Homan Z, Bricknell M. Civil-military cooperation in the early response to the covid-19 pandemic in six european countries. BMJ Mil Health 2021.

3. Pasquier P, Luft A, Gillard J, Boutonnet M, Vallet C, Pontier JM, Duron-Martinaud S, Dia A, Puyeo L, Debrus F, et al. How do we fight covid-19? Military medical actions in the war against the covid-19 pandemic in france. BMJ Mil Health 2020.

4. Pirnay JP, Selhorst P, Cochez C, Petrillo M, Claes V, Van der Beken Y, Verbeken G, Degueldre J, T’Sas F, Van den Eede G, et al. Study of a sars-cov-2 outbreak in a belgian military education and training center in maradi, niger. Viruses 2020;12(9).

5. Payne DC, Smith-Jeffcoat SE, Nowak G, Chukwuma U, Geibe JR, Hawkins RJ, Johnson JA, Thornburg NJ, Schiffer J, Weiner Z, et al. Sars-cov-2 infections and serologic responses from a sample of u.S. Navy service members - uss theodore roosevelt, april 2020. MMWR Morb Mortal Wkly Rep 2020;69(23):714–721.

6. Clifton GT, Pati R, Krammer F, Laing ED, Broder CC, Mendu DR, Simons MP, Chen HW, Sugiharto VA, Kang AD, et al. Sars-cov-2 infection risk among active duty military members deployed to a field hospital - new york city, april 2020. MMWR Morb Mortal Wkly Rep 2021;70(9):308–311.

7. Bylicki O, Paleiron N, Janvier F. An outbreak of covid-19 on an aircraft carrier. N Engl J Med 2021;384(10):976.

8. Letizia AG, Ramos I, Obla A, Goforth C, Weir DL, Ge Y, Bamman MM, Dutta J, Ellis E, Estrella L, et al. Sars-cov-2 transmission among marine recruits during quarantine. New England Journal of Medicine 2020;383(25):2407–2416.

9. US National Guard. 2020 [cited 2021 March 25]. Available from: https://www.nationalguard.mil/coronavirus/.

10. FEMA. 2021 [cited 2021 March 25]. Available from: https://www.fema.gov/fact-sheet/national-guard-deployment-extended-support-covid-19-response.

11. Mahajan S, Redlich CA, Wisnewski AV, Fazen LE, Rao LV, Kuppusamy K, Ko AI, Krumholz HM. Performance of abbott architect, ortho vitros, and euroimmun assays in detecting prior sars-cov-2 infection. medRxiv 2020:2020.2007.2029.20164343.

12. Amanat F, Stadlbauer D, Strohmeier S, Nguyen THO, Chromikova V, McMahon M, Jiang K, Arunkumar GA, Jurczyszak D, Polanco J, et al. A serological assay to detect sars-cov-2 seroconversion in humans. Nature Medicine 2020;26(7):1033–1036.

13. Statistical Atlas. 2021 [cited 2021 March 25]. Available from: https://statisticalatlas.com/state/Connecticut/Employment-Status#figure/employment-status-by-age.

14. Friedl KE, Klicka MV, King N, Marchitelli LJ, Askew EW. Effects of reduced fat intake on serum lipids in healthy young men and women at the u.S. Military academy. Mil Med 1995;160(10):527–533.

15. Mahajan S, Srinivasan R, Redlich CA, Huston SK, Anastasio KM, Cashman L, Massey DS, Dugan A, Witters D, Marlar J, et al. Seroprevalence of sars-cov-2-specific igg antibodies among adults living in connecticut: Post-infection prevalence (pip) study. Am J Med 2020.

16. Ruetalo N, Businger R, Althaus K, Fink S, Ruoff F, Hamprecht K, Flehmig B, Bakchoul T, Templin MF, Schindler M. Neutralizing antibody response in non-hospitalized sars-cov-2 patients. medRxiv 2020:2020.2008.2007.20169961.

17. Lucas C, Klein J, Sundaram M, Liu F, Wong P, Silva J, Mao T, Oh JE, Tokuyama M, Lu P, et al. Kinetics of antibody responses dictate covid-19 outcome. medRxiv 2020.

18. Price-Haywood EG, Burton J, Fort D, Seoane L. Hospitalization and mortality among black patients and white patients with covid-19. New England Journal of Medicine 2020;382(26):2534–2543.

19. Poteat T, Millett GA, Nelson LE, Beyrer C. Understanding covid-19 risks and vulnerabilities among black communities in america: The lethal force of syndemics. Annals of Epidemiology 2020;47:1–3.

20. Millett GA, Jones AT, Benkeser D, Baral S, Mercer L, Beyrer C, Honermann B, Lankiewicz E, Mena L, Crowley JS, et al. Assessing differential impacts of covid-19 on black communities. Annals of Epidemiology 2020;47:37–44.

21. Amaratunga D, Cabrera J, Ghosh D, Katehakis MN, Wang J, Wang W. Socio-economic impact on covid-19 cases and deaths and its evolution in new jersey. Ann Oper Res 2021:1–14.

