## Supplementary material for "Associations of SARS-CoV-2 serum IgG with occupation and demographics of military personnel": (Supplemental Materials Figure S1

| Item | Page # |
|---|---|
| Fig. S1. Age distribution of study subjects vs. the local population. | 2 |
| Table S1. Parsimonious adjusted logistic regression model of demographic/occupational data and SARS-CoV-2 serum IgG. | 3 |
| Table S2. Unadjusted linear regression model of demographic/occupational data and SARS-CoV-2 serum IgG. | 4 |
| Table S3. Parsimonious adjusted linear regression model of demographic/occupational data and SARS-CoV-2 serum IgG. | 5 |
| Figure S2. Spike antigen-specific IgG in serum from subjects from MOS groups with < 7 subjects/MOS code. | 6 |

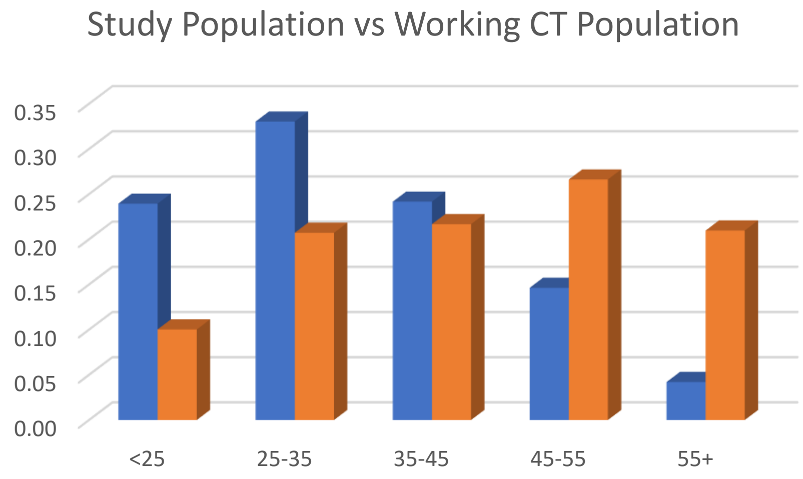


Figure S1. Age distribution of study subjects vs. the local population. The % of workers (Y-axis) in different age ranges (X-axis) are depicted. Study subjects (blue bars) data from present study and local population (orange bars) estimates are from https://statisticalatlas.com/state/Connecticut/Employment-Status#figure/employment-status-by-age.

Table S1. Parsimonious (adjusted) logistic regression model of demographic/occupational data and SARS-CoV-2 serum IgG.

| Parameter | Level | n | Spike IgG +/- | | | | | | | | |
|---|---|---|---|---|---|---|---|---|---|---|---|
| | | | Parsimonious (Adjusted) Logistic Regression Model | | | | | | | | |
| | | | Estimate | Standard Error | Odds Ratio | | | Wald Chi-Square | Pr > $\chi^2$ | Wald Chi-Square | Pr > χ2 |
| | | | | | Estimate | L95% | U95% | | | | |
| Race | Asian | 25 | 0.0921 | 0.5774 | 1.398 | 0.403 | 4.850 | 0.0255 | 0.8732 | 13.8046 | 0.0169 |
| | Black | 77 | 0.5194 | 0.3726 | 2.143 | 1.117 | 4.111 | 1.9426 | 0.1634 | | |
| | Mixed | 39 | -1.4732 | 0.8736 | 0.292 | 0.039 | 2.173 | 2.8437 | 0.0917 | | |
| | Other | 7 | 0.5437 | 0.9261 | 2.196 | 0.259 | 18.624 | 0.3446 | 0.5572 | | |
| | Unknown | 120 | 0.5610 | 0.3411 | 2.234 | 1.293 | 3.862 | 2.7048 | 0.1000 | | |
| | White | 720 | 0 (Reference) | - | 1 (Reference) | - | - | - | - | | |
| Hispanic | Hispanic | 171 | | | | | | | | | |
| | Not Hispanic | 817 | 0 (Reference) | - | 1 (Reference) | - | - | - | - | | |
| Exposed COVID | Yes | 136 | 0.3525 | 0.1323 | 2.024 | 1.205 | 3.399 | 7.1016 | 0.0077 | 7.1016 | 0.0077 |
| | Maybe/No | 852 | 0 (Reference) | - | 1 (Reference) | - | - | - | - | | |
| Number in Household | 6 or more | 67 | 0.4118 | 0.1643 | 2.279 | 1.197 | 4.339 | 6.2812 | 0.0122 | 6.2812 | 0.0122 |
| | Less than 6 | 921 | 0 (Reference) | - | 1 (Reference) | - | - | - | - | | |
| Age [years] | per year | 988 | | | | | | | | | |
| Smoking Catgory | Current | 77 | | | | | | | | | |
| | Previous | 279 | | | | | | | | | |
| | Never | 631 | 0 (Reference) | - | 1 (Reference) | - | - | - | - | | |
| Vaping Catgory | Current | 66 | | | | | | | | | |
| | Previous | 88 | | | | | | | | | |
| | Never | 834 | 0 (Reference) | - | 1 (Reference) | - | - | - | - | | |
| Alcohol | Yes - every day | 29 | | | | | | | | | |
| | Yes - weekly | 289 | | | | | | | | | |
| | Yes - monthly | 398 | | | | | | | | | |
| | No | 272 | 0 (Reference) | - | 1 (Reference) | - | - | - | - | | |
| Sex | Female | 200 | | | | | | | | | |
| | Male | 788 | 0 (Reference) | - | 1 (Reference) | - | - | - | - | | |
| Branch | Air Force | 154 | | | | | | | | | |
| | Unknown | 96 | | | | | | | | | |
| | Army | 738 | 0.0000 | . | 1 (Reference) | . | . | . | . | | |
| MOS | 2T | 7 | 0.3614 | 0.6292 | 4.095 | 0.759 | 22.102 | 0.3299 | 0.5657 | 7.3189 | 0.0257 |
| | 92F | 7 | 0.6871 | 0.6016 | 5.672 | 1.185 | 27.142 | 1.3043 | 0.2534 | | |
| | All Other | 974 | 0 (Reference) | - | 1 (Reference) | - | - | - | - | | |

Table S2. Unadjusted linear regression model of demographic/occupational data and SARS-CoV-2 serum IgG.

| Parameter | Level | n | Spike IgG ELISA OD | | | | | | | | |
|---|---|---|---|---|---|---|---|---|---|---|---|
| | | | Unadjusted Linear Regression Models | | | | | | | | |
| | | | Estimate | Standard Error | $e^{estimate}$ | L95% | U95% | t Value | Pr > \|t\| | F Value | Pr > F |
| Race | Asian | 25 | 0.0995 | 0.1740 | 1.1047 | -0.2419 | 0.4410 | 0.57 | 0.5674 | 4.98 | 0.0002 |
| | Black | 77 | 0.3382 | 0.1002 | 1.4025 | 0.1416 | 0.5349 | 3.37 | 0.0008 | | |
| | Mixed | 39 | -0.0462 | 0.1406 | 0.9549 | -0.3221 | 0.2298 | -0.33 | 0.7427 | | |
| | Other | 7 | 0.0838 | 0.3249 | 1.0874 | -0.5537 | 0.7213 | 0.26 | 0.7964 | | |
| | Unknown | 120 | 0.3331 | 0.0843 | 1.3953 | 0.1677 | 0.4986 | 3.95 | <.0001 | | |
| | White | 720 | 0 (Reference) | - | 1 (Reference) | - | - | - | - | | |
| Hispanic | Hispanic | 171 | 0.1949 | 0.0724 | 1.2151 | 0.0528 | 0.3688 | 2.69 | 0.0072 | 7.25 | 0.0072 |
| | Not Hispanic | 817 | 0 (Reference) | - | 1 (Reference) | - | - | - | - | | |
| Exposed COVID | Yes | 136 | 0.1942 | 0.0797 | 1.2143 | 0.0377 | 0.3506 | 2.44 | 0.0150 | 5.93 | 0.0150 |
| | Maybe/No | 852 | 0 (Reference) | - | 1 (Reference) | - | - | - | - | | |
| Number in Household | 6 or more | 67 | 0.2695 | 0.1092 | 1.3093 | 0.0552 | 0.4839 | 2.47 | 0.0138 | 6.09 | 0.0138 |
| | Less than 6 | 921 | 0 (Reference) | - | 1 (Reference) | - | - | - | - | | |
| Age [years] | per year | 988 | 0.0050 | 0.0026 | 1.0050 | 0.0000 | 0.0101 | 1.95 | 0.0510 | 3.82 | 0.0510 |
| Smoking Catgory | Current | 77 | 0.0409 | 0.1046 | 1.0418 | -0.1643 | 0.2461 | 0.39 | 0.6956 | 0.74 | 0.4774 |
| | Previous | 279 | -0.0660 | 0.0622 | 0.9362 | -0.1880 | 0.0561 | -1.06 | 0.2890 | | |
| | Never | 631 | 0 (Reference) | - | 1 (Reference) | - | - | - | - | | |
| Vaping Catgory | Current | 66 | 0.0684 | 0.1107 | 1.0708 | -0.1489 | 0.2857 | 0.62 | 0.5369 | 0.58 | 0.5584 |
| | Previous | 88 | 0.0907 | 0.0970 | 1.0949 | -0.0998 | 0.2811 | 0.93 | 0.3504 | | |
| | Never | 834 | 0 (Reference) | - | 1 (Reference) | - | - | - | - | | |
| Alcohol | Yes - every day | 29 | -0.0667 | 0.1691 | 0.9355 | -0.3985 | 0.2651 | -0.39 | 0.6933 | 0.85 | 0.4686 |
| | Yes - weekly | 289 | -0.0946 | 0.0730 | 0.9097 | -0.2379 | 0.0486 | -1.30 | 0.1953 | | |
| | Yes - monthly | 398 | 0.0022 | 0.0680 | 1.0022 | -0.1312 | 0.1356 | 0.03 | 0.9741 | | |
| | No | 272 | 0 (Reference) | - | 1 (Reference) | - | - | - | - | | |
| Sex | Female | 200 | -0.2648 | 0.0679 | 0.7673 | -0.3980 | -0.1317 | -3.90 | 0.0001 | 15.23 | 0.0001 |
| | Male | 788 | 0 (Reference) | - | 1 (Reference) | - | - | - | - | | |
| Branch | Air Force | 154 | -0.1314 | 0.0765 | 0.8768 | -0.2815 | 0.0186 | -1.72 | 0.0859 | 1.49 | 0.2264 |
| | Unknown | 96 | -0.0096 | 0.0928 | 0.9905 | -0.1917 | 0.1726 | -0.10 | 0.9180 | | |
| | Army | 738 | 0 (Reference) | . | 1 (Reference) | . | . | . | . | | |
| MOS | 2T | 7 | 0.9166 | 0.3259 | 2.5009 | 0.2771 | 1.5562 | 2.81 | 0.0050 | 6.49 | 0.0016 |
| | 92F | 7 | 0.7405 | 0.3259 | 2.0969 | 0.1009 | 1.3800 | 2.27 | 0.0233 | | |
| | All Other | 974 | 0 (Reference) | - | 1 (Reference) | - | - | - | - | | |

Table S3. Parsimonious (adjusted) linear regression model of demographic/occupational data and SARS-CoV-2 serum IgG.

| Parameter | Level | n | Spike IgG ELISA OD | | | | | | | | |
|---|---|---|---|---|---|---|---|---|---|---|---|
| | | | Parsimonious (Adjusted) Linear Regression Model | | | | | | | | |
| | | | Estimate | Standard Error | $e^{estimate}$ | L95% | U95% | t Value | Pr > \|t\| | F Value | Pr > F |
| Race | Asian | 25 | 0.1530 | 0.1710 | 1.1653 | -0.18 | 0.4887 | 0.89 | 0.3713 | 6.32 | <.0001 |
| | Black | 77 | 0.3893 | 0.1019 | 1.4760 | 0.19 | 0.5892 | 3.82 | 0.0001 | | |
| | Mixed | 39 | 0.0147 | 0.1382 | 1.0148 | -0.26 | 0.2859 | 0.11 | 0.9154 | | |
| | Other | 7 | 0.1256 | 0.3186 | 1.1338 | -0.50 | 0.7508 | 0.39 | 0.6936 | | |
| | Unknown | 120 | 0.3808 | 0.0837 | 1.4635 | 0.22 | 0.5452 | 4.55 | <.0001 | | |
| | White | 720 | 0 (Reference) | - | 1 (Reference) | - | - | - | - | | |
| Hispanic | Hispanic | 171 | | | | | | | | | |
| | Not Hispanic | 817 | 0 (Reference) | - | 1 (Reference) | - | - | - | - | | |
| Exposed COVID | Yes | 136 | 0.2284 | 0.0779 | 1.2566 | 0.08 | 0.3813 | 2.93 | 0.0035 | 8.59 | 0.0035 |
| | Maybe/No | 852 | 0 (Reference) | - | 1 (Reference) | - | - | - | - | | |
| Number in Household | 6 or more | 67 | 0.2375 | 0.1070 | 1.2681 | 0.03 | 0.4474 | 2.22 | 0.0266 | 4.93 | 0.0266 |
| | Less than 6 | 921 | 0 (Reference) | - | 1 (Reference) | - | - | - | - | | |
| Age [years] | per year | 988 | 0.0076 | 0.0026 | 1.0076 | 0.00 | 0.0126 | 2.94 | 0.0034 | 8.64 | 0.0034 |
| Smoking Catgory | Current | 77 | | | | | | | | | |
| | Previous | 279 | | | | | | | | | |
| | Never | 631 | 0 (Reference) | - | 1 (Reference) | - | - | - | - | | |
| Vaping Catgory | Current | 66 | | | | | | | | | |
| | Previous | 88 | | | | | | | | | |
| | Never | 834 | 0 (Reference) | - | 1 (Reference) | - | - | - | - | | |
| Alcohol | Yes - every day | 29 | | | | | | | | | |
| | Yes - weekly | 289 | | | | | | | | | |
| | Yes - monthly | 398 | | | | | | | | | |
| | No | 272 | 0 (Reference) | - | 1 (Reference) | - | - | - | - | | |
| Sex | Female | 200 | -0.2849 | 0.0671 | 0.7521 | -0.42 | -0.1532 | -4.25 | <.0001 | 18.04 | <.0001 |
| | Male | 788 | 0 (Reference) | - | 1 (Reference) | - | - | - | - | | |
| Branch | Air Force | 154 | | | | | | | | | |
| | Unknown | 96 | | | | | | | | | |
| | Army | 738 | 0.0000 | - | 1 (Reference) | - | - | - | - | | |
| MOS | 2T | 7 | 0.9565 | 0.3186 | 2.6025 | 0.33 | 1.5817 | 3.00 | 0.0027 | 6.50 | 0.0016 |
| | 92F | 7 | 0.6412 | 0.3188 | 1.8988 | 0.02 | 1.2668 | 2.01 | 0.0445 | | |
| | All Other | 974 | 0 (Reference) | - | 1 (Reference) | - | - | - | - | | |

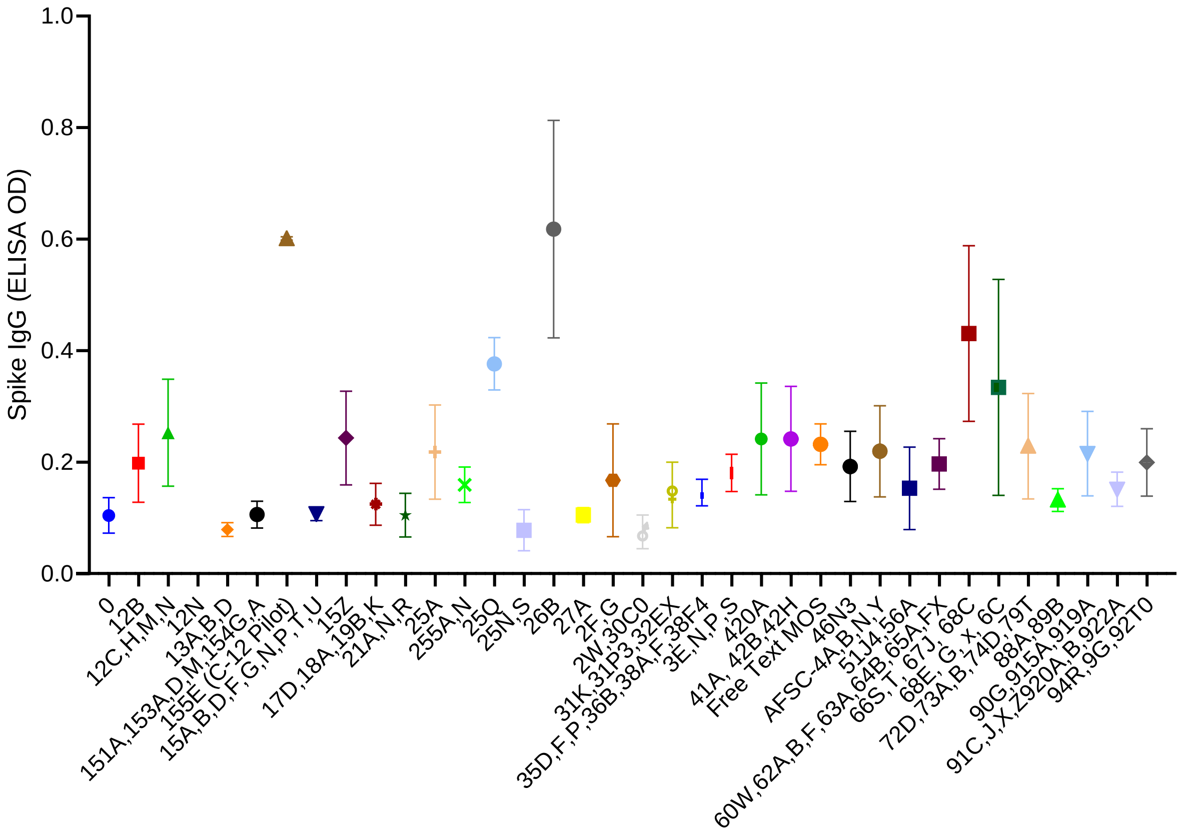


Figure S2. Spike antigen-specific IgG in serum from subjects with different MOS codes. The mean and standard error of spike IgG ELISA OD values are shown for subjects that belong to different MOS groups with N ≤ 6 per MOS.
